# Prevalence and Severity of Nicotine Dependence in India: A Systematic Review and Meta-Analysis Protocol

**DOI:** 10.1101/2025.09.28.25336830

**Authors:** Chandru Sivamani, S Kavya, Lavanya Ayyasamy, Revathy Ajayan, S Uvaraja, Mamta Mor, Sombuddha Biswas, Kavipriya Outtamane, Sooriyaprasannan Sriram

**Author notes:** **Corresponding author/First author** Dr Chandru Sivamani, BDS, MPH, Joint Secretary, Care Max Foundation, Puducherry 605009, India. **Co-author details 1. Dr Kavya S, BDS, MPH,** Member,;, **2. Lavanya Ayyasamy, BSc, MSc (Epidemiology and Biostatics),**;, **3. Dr Revathy Ajayan, BDS, MPH,**;, **4. Dr. S Uvaraja, BDS, MDS,** Assistant Professor, **5. Dr Mamta Mor, BDS, MPH,** Member, **6. Dr Sombuddha Biswas, BDS, MPH,** Member, **7. Dr Kavipriya Outtamane, BDS, MBA, MPH,** Chief Dental Surgeon (NFSG), **8. Dr Sooriyaprasannan Sriram, MBBS, PGDCD,** President.

## Abstract

**Background:** Despite the high prevalence on tobacco use in India, evidence on nicotine dependence and its severity across the demographic groups is limited. Therefore, this review aims to estimate the pooled prevalence and severity of nicotine dependence among tobacco users in India, stratified by age, gender, type of tobacco, and rural–urban setting.

**Methods:** The protocol has been prospectively registered in PROSPERO (ID: CRD420251133993) and adheres to the PRISMA-P 2015 guidelines. Comprehensive literature searches will be conducted across Medline, Cochrane Library, Embase, Scopus, and grey literature sources. Eligible studies will include community-based research reporting nicotine dependence assessed using the Fagerström Test for Nicotine Dependence (FTND) or FTND-ST for smokeless tobacco. Two independent reviewers will carry out study selection, data extraction, and risk-of-bias assessment using the JBI checklist and the Newcastle–Ottawa Scale. Pooled prevalence will be calculated using a random-effects model, supplemented by subgroup analyses, sensitivity analyses, and meta-regression. Publication bias will be assessed through Deviation from Expected Inverse Variance (DOI) plots for graphical asymmetry evaluation or by funnel plots, alongside the Luis Furuya–Kanamori (LFK) index, where values within ±1 suggest no asymmetry, ±1 to ±2 indicate minor asymmetry, and >±2 represent major asymmetry. The overall certainty of evidence will be appraised using the GRADE framework.

**Discussion:** By generating robust estimates of nicotine dependence and its severity among tobacco users in India, this review will address a significant evidence gap in the field. The findings will be valuable for policymakers, public health professionals, and clinicians, enabling them to design context-specific tobacco control strategies, enhance cessation services, and develop targeted interventions. Such measures will contribute meaningfully to achieving Sustainable Development Goals 3 and 3.a, which aim to reduce premature mortality from non-communicable diseases.

## Introduction

Globally, the tobacco epidemic remains most significant public health problem causing more than 8 million deaths annually and also the exposure to second-hand smoke contributing approximately 1.2 million deaths in every year.[1] The use of tobacco causing more than 20 different types of cancers (including cancers of the lung, oral cavity, pharynx, larynx, esophagus, stomach, pancreas, liver, kidney, bladder and cervix) and certain hematologic malignancies such as acute myeloid leukemia.[2] In addition to cancer, it is a major risk factor for several chronic conditions such as cardiovascular diseases (coronary heart disease, stroke, and peripheral arterial disease), chronic respiratory diseases (COPD, chronic bronchitis, and emphysema), type 2 diabetes, adverse reproductive outcomes, and immune dysfunction.[2] The majority of tobacco-attributable deaths are in the low- and middle-income countries, where the tobacco industry frequently engages in the intensive marketing strategy and interference.[1]

In India, around 36.8% of the population are current users of tobacco in any form, and tobacco consumption accounts for approximately 1.35 million deaths, representing 17.8% of all deaths in the country, and accounts for nearly 28.9 million disability-adjusted life years (DALYs) lost in 2021.[3,4] Unlike many other countries where smoked tobacco predominates, India has a higher consumption of smokeless tobacco, including products such as khaini, gutkha, zarda, and betel quid, which are deeply embedded in cultural practices.[5] Evidence from community as well as facility-based studies reported that the nicotine dependence in India ranges from 37% to 75% across different populations.[6–8] However, there is limited nationally representative evidence on nicotine dependence and its severity by demographic subgroups, tobacco type, and setting (rural-urban) highlighting a critical knowledge gap.

India has made significant progress in tobacco control through the Cigarettes and Other Tobacco Products Act (COTPA, 2003), which regulates the production, trade, and sale of tobacco products, prohibits advertising, mandates large pictorial health warnings, and restricts smoking in public places and near educational institutions.[9] Ratification of the WHO Framework Convention on Tobacco Control (FCTC) in 2004, establishment of tobacco cessation clinics since 2002, the 2019 ban on electronic cigarettes and heated tobacco products, and fiscal measures such as Good Service Tax (GST) based taxation (28%) further strengthen the regulatory framework.[1,4,10] Despite these measures, enforcement gaps, limited access to cessation services, and the cultural acceptability of smokeless tobacco limit effectiveness. These contextual challenges indicate that a one-size-fits-all approach may be insufficient to address the heterogeneous patterns of nicotine dependence observed across India.

Given these complexities, systematically documenting the prevalence and severity of nicotine dependence is critical. Therefore, this study aims to estimate the pooled prevalence of nicotine dependence among tobacco users in India and to assess the severity of dependence, to estimate the pooled mean scores of FTND/FTND-ST scales, and subgroup variations by age, sex, tobacco form (smoking and smokeless) and setting (rural and urban). Such evidence will inform context-sensitive interventions, strengthen public health programs, and guide policymakers toward achieving SDG 3 and 3.a, emphasizing reduction of premature mortality from non-communicable diseases.[11]

## Materials and methods

This review on the prevalence and severity of nicotine dependence in India was prospectively registered in PROSPERO on 26^th^ August 2025 (ID: **CRD420251133993**) and any required amendments will be reflected in the PROSPERO record. The protocol has been developed in adherence with the Preferred Reporting Items for Systematic Review and Meta-Analysis Protocols (PRISMA-P) 2015 statement (Supplementary file 1).[12] This SRMA will also be conducted in accordance with the relevant chapter of the Joanna Briggs Institute (JBI) manual for evidence synthesis.[13]

### Eligibility criteria

The inclusion criteria will be guided by the CoCoPop framework (Condition–Context– Population). In this review, the condition is defined as nicotine dependence, assessed using either the Fagerström Test for Nicotine Dependence (FTND) for smoked tobacco or the FTND-ST for smokeless tobacco. Eligible studies must report community-based prevalence data and focus on the Indian population, irrespective of age and gender. There will be no restriction on the year of publication; however, only studies published in English will be considered. Studies focusing on facility-based settings, case series, case reports, case-control studies, commentaries, reviews and letters to the editor will be excluded from this review.

### Search strategy

We will conduct comprehensive literature searches across multiple databases, including Medline (via PubMed), Cochrane Library, Embase, and Scopus, along with grey literature sources. The initial search will be carried out on October 1, 2025, and updated in January 2026 prior to the final analysis. Search strategies will be structured following the CoCoPop framework by combining relevant controlled vocabulary (MeSH/EMTREE) with free-text keywords, including “tobacco dependence,” “nicotine dependence,” “FTND,” “FTND-ST,” “smoked tobacco,” “smokeless tobacco,” “India,” and “community-based,” adapted for each database functionality (Supplementary File 2). Boolean operators such as AND/OR will be applied to combine terms appropriately.

### Data management

The search results from all databases will be exported in RIS/nbib format and imported into Rayyan, a web-based tool for screening and managing systematic review records.[14] Duplicate entries will be identified and manually removed prior to screening. Independent title and abstract screening will be conducted in Rayyan by two reviewers, with disagreements resolved by a third reviewer. Records classified as ‘include’ or ‘maybe’ will proceed to full-text screening. Final inclusions will be exported to Microsoft Excel for data extraction and analyzed using R Studio (version 4.4.2). The certainty of evidence will be evaluated using the Grading of Recommendations, Assessment, Development and Evaluation (GRADE) approach, and the findings will be summarized in tables generated through the GRADEpro web application (https://www.gradepro.org/).[15]

### Data extraction

Following full-text review, data will be extracted from studies that fulfils the eligibility criteria. To ensure consistency, we will use a standardized data extraction form, adapted from JBI manual.[13] Data will be extracted from each eligible study across three key areas. This will include bibliographic and general information such as author, title of the study, year of publication, and location. Methodological details will also be recorded, covering study design, setting, population characteristics, inclusion and exclusion criteria, sample size, and type of tobacco use. Additionally, outcome data will be collected, comprising the prevalence and severity of nicotine dependence reported as proportions or means (SD), along with subgroup analyses by age, gender, duration of smoking, region, and rural or urban status.

### Assessment of Risk of Bias

The methodological quality and risk of bias of included studies will be assessed using a dual approach: for prevalence and severity of nicotine, the JBI critical appraisal checklist will be applied (**Table 1**).[13] For continuous outcomes, such as mean FTND or FTND-ST scores, and for comparative analyses, the Newcastle–Ottawa Scale (NOS) will be used to assess study quality.[16] The NOS evaluates three domains: Selection, Comparability, and Outcome/Exposure, with a maximum score of nine stars. Studies attaining seven or more stars will be classified as high quality, those scoring five to six stars as moderate quality, and those with four or fewer stars as low quality. Two reviewers will independently conduct all assessments and the discrepancies resolved through discussion or consultation with a third reviewer.

**Table 1:**
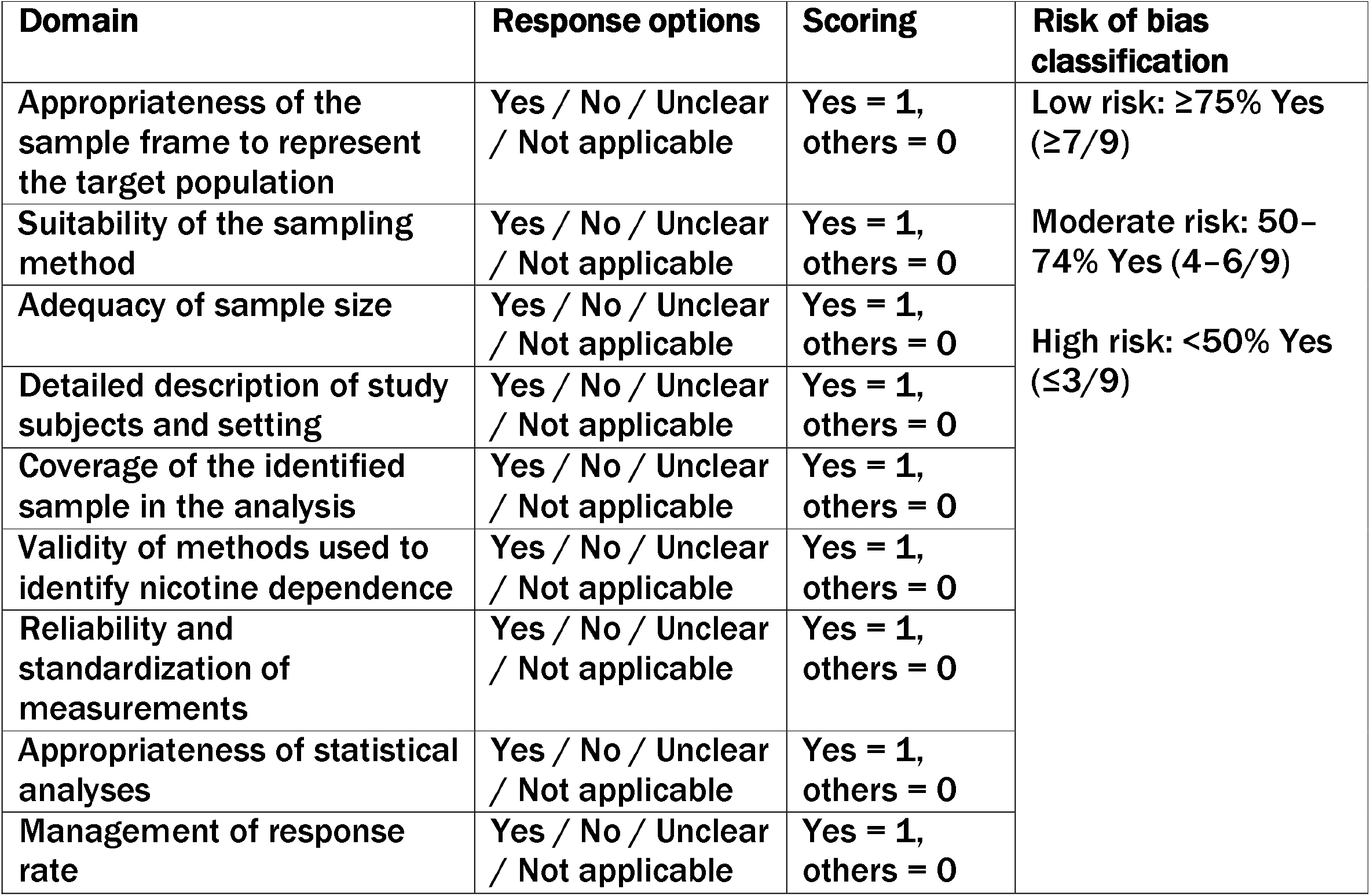
Assessment of risk of bias.

### Data synthesis

A comprehensive table summarizing the key characteristics and findings of the included studies will be developed in accordance with the eligibility criteria to clearly present essential information for each study. This will be accompanied by a narrative synthesis to integrate and interpret the findings, and a meta-analysis to compute pooled estimates of the prevalence and severity of nicotine dependence. The prevalence of nicotine dependence will be pooled using random-effects model with 95%CI (Confidence Interval). Subgroup analyses will be performed by age group (13–24, 25– 44, ≥45 years), in accordance with GYTS-Global Youth Tobacco Survey and GATS-Global Adult Tobacco Survey)[17,18], sex, geographic region, and setting (rural versus urban) when at least two studies are available per subgroup. Continuous outcomes, including mean FTND scores for smoked tobacco users and mean FTND-ST scores for smokeless tobacco users, will be pooled separately using inverse-variance random-effects model (with 95% CI). Standard deviations not reported in studies will be derived from standard errors, CIs, or interquartile ranges as needed. Comparative analyses between smoked and smokeless tobacco users will use mean differences if reported within the same study; otherwise, descriptive comparisons will be provided.

### Meta-bias(es)

Statistical heterogeneity will be evaluated using Cochran’s Q test and Higgins’ I^2^ statistic. [19] In cases where ten or more studies are available, publication bias will be assessed using DOI (Deviation from Expected Inverse Variance) plots, which provide improved sensitivity over traditional funnel plots.[20] Asymmetry will be evaluated using the Luis Furuya-Kanamori (LFK) index, with an LFK value within the range of ±1 indicating absence of asymmetry, values between ±1 and ±2 representing minor asymmetry, and values exceeding ±2 denoting substantial asymmetry. Forest plots will be generated to display pooled prevalence estimates, mean FTND and FTND-ST scores, and comparative analyses between smoked and smokeless tobacco users. Severity distributions (mild, moderate, severe dependence) will be visualized using stacked bar charts or separate forest plots, allowing clear comparison across studies. All plots will include appropriate CIs, weights, and study-level estimates, and will be interpreted in conjunction with LFK index scores to evaluate potential bias and heterogeneity in the meta-analysis and also the robustness of the pooled prevalence estimates will be assessed (GRADE).

## Discussion

This systematic review and meta-analysis protocol is designed to provide comprehensive evidence on the prevalence and severity of nicotine dependence among tobacco users in India. While numerous studies have examined patterns of tobacco use, there is no consolidated national-level synthesis that accounts for variations by age, sex, type of tobacco product, and rural–urban setting. By systematically pooling estimates from community-based studies, this review will address an important knowledge gap and generate evidence that can directly inform tobacco control policies and cessation strategies.

A major strength of this protocol lies in the use of standardized and validated assessment tools, namely the FTND for smoked tobacco and the FTND-ST for smokeless tobacco. This will enhance comparability across studies and provide more robust estimates of dependence levels. Furthermore, by applying the CoCoPop framework and following established guidelines such as PRISMA-P and the JBI manual, the review ensures methodological rigor. The dual approach to risk of bias assessment using the JBI checklist and NOS, combined with the GRADE approach for certainty of evidence, further strengthens the reliability of the findings.

Nonetheless, several limitations are anticipated. Restricting inclusion to studies published in English may exclude relevant data available in regional languages. Considerable heterogeneity is expected due to differences in study designs, populations, and cultural contexts of tobacco use across India. While subgroup analyses and random-effects models will help account for this variability, residual heterogeneity may persist. In addition, reliance on self-reported measures of tobacco use and nicotine dependence may introduce reporting bias.

Despite these challenges, the findings of this review will have significant implications. Generating pooled estimates of nicotine dependence and severity across diverse subgroups will provide policymakers with evidence to tailor interventions, such as designing culturally sensitive cessation programs and prioritizing high-burden groups. This work will also support India’s commitments under the WHO Framework Convention on Tobacco Control (FCTC) and Sustainable Development Goals (SDG 3.4 and 3.a), particularly in reducing premature mortality from non-communicable diseases.[4,11]

In conclusion, this systematic review and meta-analysis will generate timely and policy-relevant evidence on nicotine dependence in India. By bridging the existing evidence gaps, it has the potential to inform targeted tobacco control strategies, strengthen cessation services, and contribute to reducing the health and economic burden associated with tobacco use.

## Data Availability

As this is a study protocol, data availability is not applicable at this stage. However, upon completion of data extraction, the extracted dataset and the codebook used for analysis will be made available upon reasonable request.

## Author contributions

**Conceptualization:** Chandru Sivamani (CS), Kavya S (KS), Lavanya Ayyasamy (LA), Revathy Ajayan (RA), Mamta Mor (MM), Sombuddha Biswas (SB), Kavipriya Outtamane (KO), Uvaraja S (US), and Sooriyaprasannan Sriram (CS); **Methodology:** CS and KS; **Writing – Original Draft:** CS, KS, MM and RA; **Writing – Review & Editing:** CS, KS, LA, RA, MM, SB, KO, US and SS; All authors approved the final version of the manuscript, and agree to be accountable for all aspects of the work. Chandru Sivamani acts as the **guarantor** for this work.

## Funding

This study did not receive financial support from any governmental, commercial, or non-profit funding bodies.

## Declarations

### Ethical approval and consent to participate

As this study is a secondary analysis based on previously published literature, ethical approval and participant consent are not applicable.

### Conflicts

The authors state that there are no conflicts of interest related to this work.

## Notes

### Competing Interest Statement

The authors have declared no competing interest.

### Clinical Protocols

https://www.crd.york.ac.uk/PROSPERO/view/CRD420251133993.

### Funding Statement

This study did not receive any funding

