## Supplementray file 2 for "Prevalence and Severity of Nicotine Dependence in India: A Systematic Review and Meta-Analysis Protocol"

Supplementary file 2: Search Strategy for all the database

PubMed

((((("nicotine dependence"[Title/Abstract]) OR ("tobacco dependence"[Title/Abstract])) OR ("nicotine addiction"[Title/Abstract])) OR ("tobacco addiction"[Title/Abstract])) OR (((((((((((("fagerström test"[Title/Abstract]) OR ("fagerstrom test for nicotine dependence"[Title/Abstract])) OR (FTND score[Title/Abstract])) OR (FTND[Title/Abstract])) OR ("fagerstrom nicotine dependence scale"[Title/Abstract])) OR ("smokeless tobacco dependence scales"[Title/Abstract])) OR (FTND-ST[Title/Abstract])) OR ("oklahoma scale for smokeless tobacco dependence"[Title/Abstract])) OR (OSSTD[Title/Abstract]) OR ("tobacco dependence screener"[Title/Abstract])) OR (TDS[Title/Abstract])) OR (SSTDS[Title/Abstract])) OR ("nicotine severity"[Title/Abstract]))) AND (((india [MeSH Terms]) OR (india[Title/Abstract])) OR (indian[Title/Abstract]))

Scopus

TITLE-ABS-KEY (("nicotine dependence") OR ("tobacco dependence") OR ("nicotine addiction") OR ("tobacco addiction") OR ("fagerström test") OR ("fagerstrom test for nicotine dependence") OR ("FTND score") OR (FTND) OR ("fagerstrom nicotine dependence scale") OR ("smokeless tobacco dependence scales") OR (FTND-ST) OR ("oklahoma scale for smokeless tobacco dependence") OR (OSSTD) OR ("tobacco dependence screener") OR (TDS) OR (SSTDS) OR ("nicotine severity")) AND ((india) OR (indian)))

Embase

('nicotine dependence':ti,ab OR 'tobacco dependence':ti,ab OR 'nicotine addiction':ti,ab OR 'tobacco addiction':ti,ab OR 'fagerström test':ti,ab OR 'fagerstrom test for nicotine dependence':ti,ab OR 'FTND score':ti,ab OR 'FTND':ti,ab OR 'fagerstrom nicotine dependence scale':ti,ab OR 'smokeless tobacco dependence scales':ti,ab OR 'FTND-ST':ti,ab OR 'oklahoma scale for smokeless tobacco dependence':ti,ab OR 'OSSTD':ti,ab OR 'tobacco dependence screener':ti,ab OR 'TDS':ti,ab OR 'SSTDS':ti,ab OR 'nicotine severity':ti,ab) AND ('india':ti,ab OR 'indian':ti,ab OR 'India'/exp)

Cochrane

([Title/Abstract] "nicotine dependence" OR [Title/Abstract] "tobacco dependence" OR [Title/Abstract] "nicotine addiction" OR [Title/Abstract] "tobacco addiction" OR [Title/Abstract] "fagerström test" OR [Title/Abstract] "fagerstrom test for nicotine dependence" OR [Title/Abstract] "FTND score" OR [Title/Abstract] "FTND" OR [Title/Abstract] "fagerstrom nicotine dependence scale" OR [Title/Abstract] "smokeless tobacco dependence scales" OR [Title/Abstract] "FTND-ST" OR [Title/Abstract] "oklahoma scale for smokeless tobacco dependence" OR [Title/Abstract] "OSSTD" OR [Title/Abstract] "tobacco dependence screener" OR [Title/Abstract] "TDS" OR [Title/Abstract] "SSTDS" OR [Title/Abstract] "nicotine severity") AND ([MeSH] "India" OR [Title/Abstract] "India" OR [Title/Abstract] "Indian")
